# Disparities in Cardiovascular Disease in Women with Adverse Pregnancy Outcomes

**DOI:** 10.1101/2025.09.03.25335013

**Authors:** Maria Natalia Chaves Rivera, Santiago Clocchiatti-Tuozzo, Daniela Renedo, Cyprien A. Rivier, Shufan Huo, Ryan M. Hebert, Mitchell S.V. Elkind, Aaron Lazorwitz, Charles C. Matouk, Reshef Tal, Guido J Falcone

## Abstract

**Background:** Although adverse pregnancy outcomes (APOs) have been linked to increased cardiovascular disease (CVD) risk, most studies have focused on homogenous populations. We tested the hypothesis that APOs and race and ethnicity synergistically increase the risk of CVD.

**Methods:** We conducted retrospective analyses of data collected by *All of Us*, a large and diverse population study in the United States. We included women with a single lifetime delivery and no history of CVD before that delivery. Our exposures of interest were APOs (defined as any preeclampsia, eclampsia, gestational hypertension and diabetes, preterm delivery, fetal growth restriction, or placental abruption) and self-reported race and ethnicity. Our primary outcome was incident CVD ≥1 year postpartum, defined as any stroke (ischemic or hemorrhagic), myocardial infarction, or heart failure. Multivariable Cox proportional hazard models estimated the association between APO, race and ethnicity and CVD, and product terms were used to test for synergistic contributions (interaction) between APO and race and ethnicity.

**Results:** We included 10,760 women in our analysis, who had a mean (SD) age of 30 (±6) years at first delivery. Of these, 4,204 (39%) had at least one APO. After a median (interquartile range) follow-up of 7.02 (3.82-13.26) years, 217 (5.2%) women with APO sustained CVD, while only 252 (3.8%) without APO did (*p*<0.001). Multivariable Cox regressions confirmed these results, showing that women with APO had a significantly increased risk of CVD (HR: 1.45; 95%CI: 1.17-1.80). Race and ethnicity significantly modified the association between APO and CVD (interaction-*p*=0.003): compared with White women without APO, Black women with APO had a four-fold higher risk of CVD (HR: 4.04; 95%CI: 2.81–5.79), which was also substantially higher than the risk observed in White women with APO (HR: 2.01; 95%CI: 1.43–2.84).

**Conclusions:** In a large and diverse cohort of American women with a single lifetime delivery, APOs were associated with an increased risk of CVD, and these risks were even greater among Black women. These findings highlight the potential to integrate reproductive health history and non-medical determinants of health to enhance CVD prevention and health equity using personalized medicine. Further research is needed to identify the mechanisms that lead to synergistic contributions by APOs and race and ethnicity.

## INTRODUCTION

Adverse pregnancy outcomes (APOs), including hypertensive disorders of pregnancy (preeclampsia, eclampsia and gestational hypertension), gestational diabetes, preterm delivery, fetal growth restriction, and placental abruption, have increasingly been recognized as important indicators of long-term cardiovascular disease (CVD) risk in women.^1^ Despite improvements in maternal care, APOs affect a substantial proportion of pregnancies in the United States, with long-term implications for women’s health that extend well beyond the postpartum period.^1^ Growing evidence suggests that pregnancy may serve as an early “stress test,” revealing underlying vascular or metabolic susceptibilities that predispose women to future CVD.^2^

Recognizing this, the American Heart Association released a 2021 scientific statement underscoring the importance of incorporating APO history into CVD risk assessment for women.^1^ That statement highlighted two critical priorities: the scarcity of data linking APOs to CVD in diverse populations, and the urgent need for health system changes to better address the unique cardiovascular needs of women.^3^

Yet, much of the existing research originates from relatively homogeneous populations, limiting the generalizability of findings across the diverse racial and ethnic groups that comprise the modern U.S. population.^1^ Disparities in CVD outcomes between racial and ethnic groups are well documented, particularly for people self-identifying as Black, yet few studies have examined whether the excess risk conferred by APOs varies across these groups.^1,4^ Addressing these knowledge gaps is essential to advance equitable risk stratification and intervention strategies that incorporate non-medical determinants of health, paving the way toward more personalized approaches to CVD prevention.

In this study, we leveraged data from *All of Us*, a large and diverse population study in the US, to investigate the association between APOs and CVD among women with a single lifetime delivery. We tested the hypothesis that APOs and race and ethnicity synergistically increase the risk of CVD.

## METHODS

### Study Design and Participants

We conducted a longitudinal analysis of prospectively collected data within *All of Us* to examine the associations between APOs, race and ethnicity and CVD. The National Institutes of Health’s *All of Us* is a large ongoing prospective cohort study within the United States that aims to enroll 1 million Americans aged 18 and older.^5^ *All of Us* welcomes participants from all backgrounds to reduce disparities in medical research. Among persons who consent, *All of Us* collects survey questionnaires capturing detailed past and present medical history data, electronic health records, physical measurements (e.g.: blood pressure, height, weight). More information regarding *All of Us*, its study design, and logistics can be found elsewhere.^6^ Inclusion criteria for the current analyses were women with a single, lifetime, electronic health record-recorded birth (Table S1), without any prevalent CVD. Prior CVD was defined as any stroke (ischemic or hemorrhagic), myocardial infarction or heart failure before first delivery. We restricted to women with a single lifetime delivery to ensure fixed exposure status, in turn avoiding misclassification that could occur if women without an APO in their first pregnancy later developed an APO in subsequent pregnancies, which could bias time-to-event analyses and violate proportional hazards assumptions. The *All of Us* institutional review board approved the protocol and study (https://allofus.nih.gov/about/who-we-are/institutional-review-board-irb-of-all-of-us-research-program), and all participants or their legally designated surrogates provided written informed consent. We followed the Strengthening the Reporting of Observational Studies in Epidemiology (STROBE) reporting guidelines.^7^

### Ascertainment of Exposures: Adverse Pregnancy Outcomes and Race and Ethnicity

Our first exposure of interest was APO, defined as the presence/absence of one or more of the following conditions during pregnancy, for which the body of evidence is strong in their association with CVD events in later life:^1^ preeclampsia, eclampsia, gestational hypertension, gestational diabetes, preterm delivery, fetal growth restriction, or placental abruption. These were ascertained using International Classification of Diseases Codes 9^th^ and 10^th^ edition (ICD 9 and 10) or Systematized Nomenclature of Medicine Clinical Terms (SNOMED) codes (Table S2).^8,9^ Our second exposure of interest was race and ethnicity, which was a self-reported variable, collected via survey questionnaire at enrollment, and defined as the following categories: White, Asian, Hispanic, Black or Other/Multiethnic.

### Ascertainment of Outcomes

The primary outcome of interest was CVD, defined as a composite of any incident ischemic stroke, hemorrhagic stroke, myocardial infarction, or heart failure, identified using ICD-9 and - 10 and SNOMED codes (Table S3). Secondary outcomes were each component of CVD analyzed separately.

### Covariates

Smoking was ascertained via questionnaire data. Overweight and obesity were calculated using participants’ body mass index (≥25 and ≥30, respectively). Body mass index was calculated as weight in kilograms divided by height in meters squared by the *All of Us* core team using height and weight measured at study enrollment. Hypertension, diabetes mellitus, hyperlipidemia, and atrial fibrillation were ascertained from individual-level electronic health records, clinical notes, and self-report data using ICD-9 and −-10, and SNOMED codes (Table S4).

### Statistical Analysis

Discrete variables are presented as counts (%), and continuous variables as means (standard deviation [SD]) or medians (interquartile range [IQR]), as appropriate. To protect participants from re-identification and to comply with *All of Us* Data Statistics and Dissemination policy, cell counts <20 are not reported. Time-to-event analyses were used to estimate the association between APOs and outcomes, starting at 1 year postpartum to avoid the immediate postpartum period, which is known to carry elevated CVD risk.^10,11^ Participants were followed until the outcome of interest, death, or 25 years after the 1-year postpartum date, at which point >90% of participants had complete follow-up data.

#### Primary Analyses

We first plotted Kaplan–Meier curves to visualize unadjusted associations between APO (present vs. absent) and the primary outcome of CVD, with statistical comparison using the log-rank test. We then fit multivariable Cox proportional hazards models to estimate hazard ratios (HRs) and 95% confidence intervals (CI) for APO and CVD. We used two hierarchical models for our multivariable Cox regressions. Model 1 adjusted for age, race and ethnicity. Model 2 adjusted for model 1 covariates plus cardiovascular risk factors and comorbidities (hypertension, diabetes mellitus, hyperlipidemia, smoking, overweight/obesity, and atrial fibrillation) known to associate as potential confounders with our outcomes of interest. ^12–17^ We used a complete case analysis, excluding participants with missing data for the exposure, outcome, or covariates. The proportional hazards assumption was evaluated using log–log survival plots. We used product terms to assess formal statistical interaction between APO and race and ethnicity for the primary outcome. When the interaction was statistically significant, we performed Cox proportional hazards models to estimate hazard ratios for APO within each race and ethnicity category.

#### Secondary Analyses

For our secondary outcomes (ischemic stroke, hemorrhagic stroke, myocardial infarction, and heart failure) we applied the same time-to-event framework as in the primary analysis, including the same covariate adjustment strategy.

#### Sensitivity Analyses

We conducted three separate sensitivity analyses, repeating the primary analysis framework with specific caveats. First, we used an expanded cohort including all women with ≥1 delivery (n=14,749) and no prior CVD. Because APO status could change across pregnancies and CVD events could occur before the onset of an APO in later pregnancies, time-to-event methods were not applied. Instead, we used multivariable logistic regressions to evaluate the cross-sectional association between ever experiencing an APO (in any pregnancy) and CVD >1 year postpartum and used product terms to assess for formal statistical interaction between APO and race and ethnicity, adjusting for the same covariates as in the primary analysis. Second, we used the complete follow-up period available after the first recorded delivery (∼43 years). Third, we conducted time-to-event analysis starting from the day of delivery, that is, including the 1 year after delivery.

#### Exploratory Analyses

Finally, we conducted two exploratory analyses. First, we fit multivariable linear regressions adjusting for the same covariates as in the primary analysis, to assess how a history of APO associates with the age at which women sustain CVD. Second, because the magnitude and mechanisms of CVD risk may vary across individual APOs, we analyzed each APO condition separately fitting multivariable Cox proportional hazard models adjusting for the same covariates as in the primary analysis, to assess which individual APO was driving the association between any APO and CVD.

All analyses were conducted from February 2025 to August 2025 using the R statistical package version 4.5 (R Core Team, https://www.R-project.org/)

### Data Availability

*All of Us* data are publicly available at www.allofus.nih.gov. We used the release version 8, which includes data from all participants who enrolled in the program’s inception on May 30, 2017, to October 1, 2023. All data access and analyses were conducted within a secure informatics workspace provided by the National Institutes of Health.

## RESULTS

Among the total 633,547 AoU participants, 395,987 (62.5%) were women. Of these, 10,760 (2.72%) had one single recorded delivery with no prior history of CVD and were thus included in the final study cohort (Figure 1). The mean (standard deviation [SD]) age at first delivery was 30 (±6) years, and 4,204 (39%) women had one or more APOs while 6,556 (61%) did not. Table S4 reports the distribution of individual APOs sustained, showing that gestational hypertension (1,557, [26%]), preterm birth (1,264 [21.08%]), and gestational diabetes (1,056 [17.61%]) were the most prevalent APOs, respectively. Women with APO were more likely to self-identify as Black, have a higher prevalence of cardiovascular risk factors and higher physical measurements such as systolic blood pressure, hemoglobin A1c and body mass index (Table 1). The mean yearly income and deprivation index were similar among both groups.

**Figure 1.**
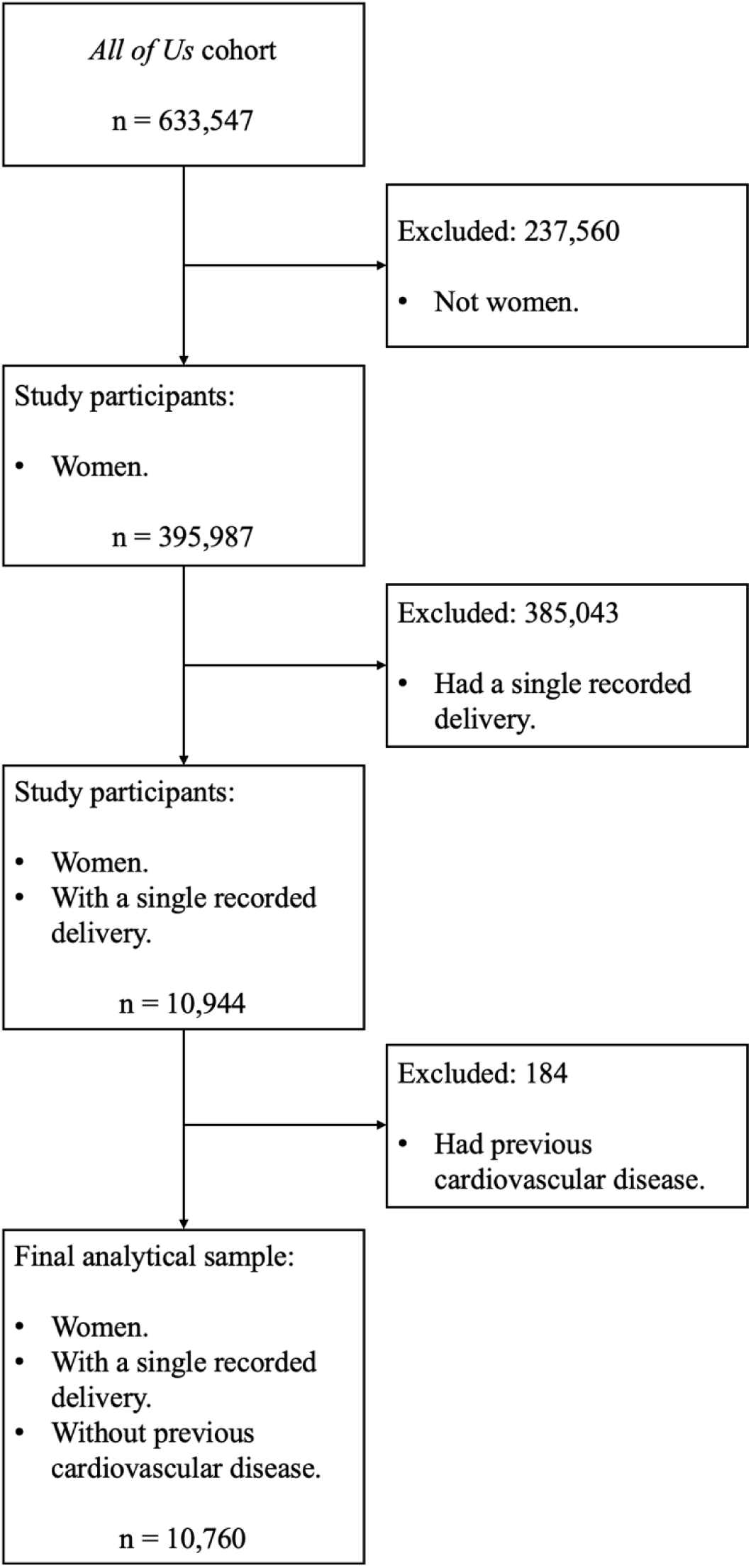
Flowchart showing the steps taken to arrive at the final analytical sample. Cardiovascular disease was defined as any ischemic stroke, intracranial hemorrhage, myocardial infarction, or heart failure.

**Table 1.**
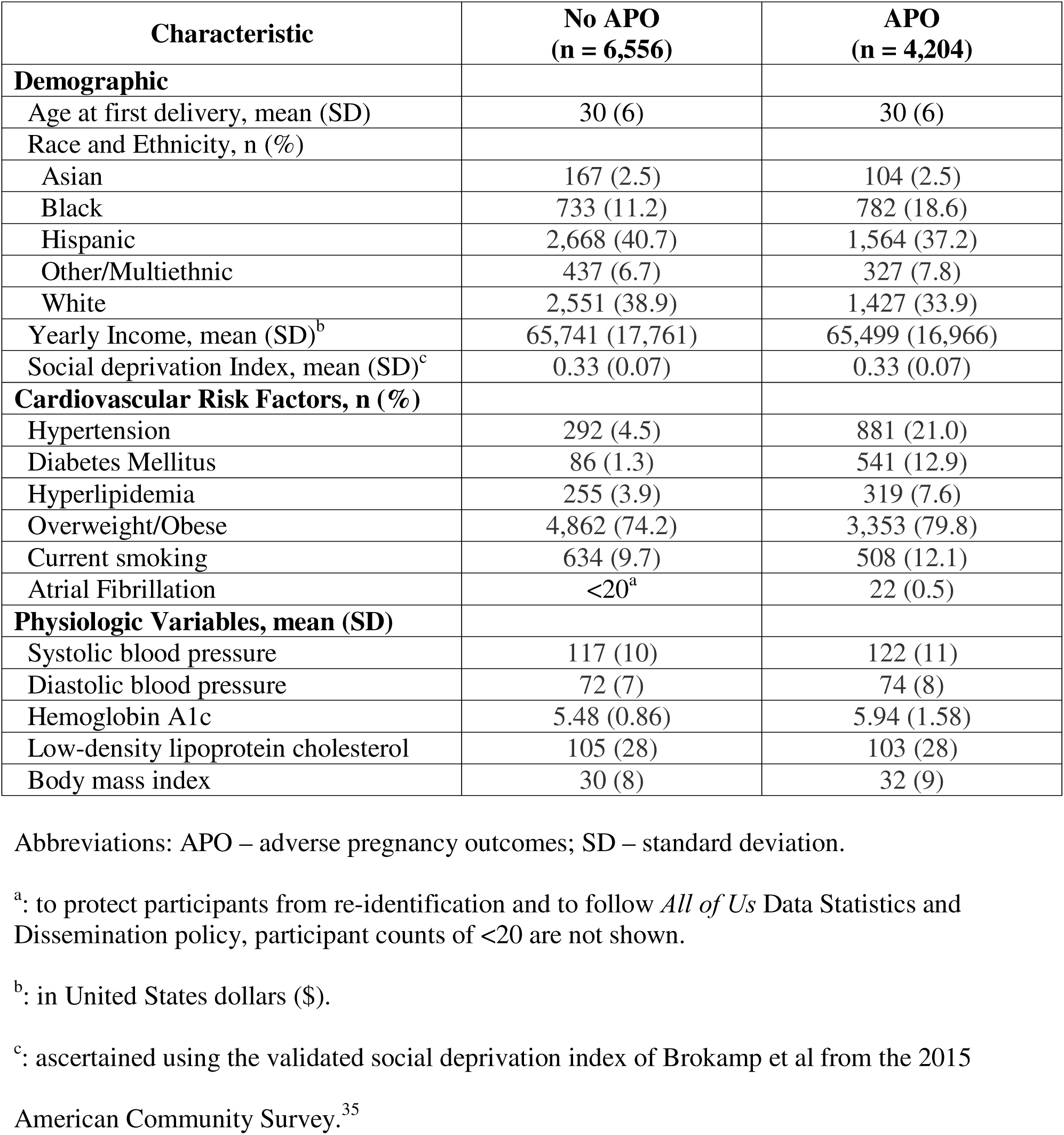
Baseline characteristics of the study cohort.

### Primary Analyses: Adverse Pregnancy Outcomes and Cardiovascular Disease and the Synergistic Effects of Race and Ethnicity

After a median follow-up time of 7.02 years (interquartile range [IQR], 3.82–13.26 years), 423 women (3.93%) experienced CVD, including 208 (4.9%) women with a history of APO and 215 (3.3%) without (Figure 2; log-rank *p*<0.001). In fully adjusted Cox proportional hazards models, a history of APO was associated with a 45% increased hazard of CVD compared to no APO history (HR: 1.45; 95%CI: 1.17–1.80; Table 2). Results were consistent across covariate-adjusted models (Table 2). Visual inspection of complementary log-log survival plots demonstrated approximate parallelism between groups (Figure S2), further supporting the validity of the proportional hazards assumption.

**Figure 2.**
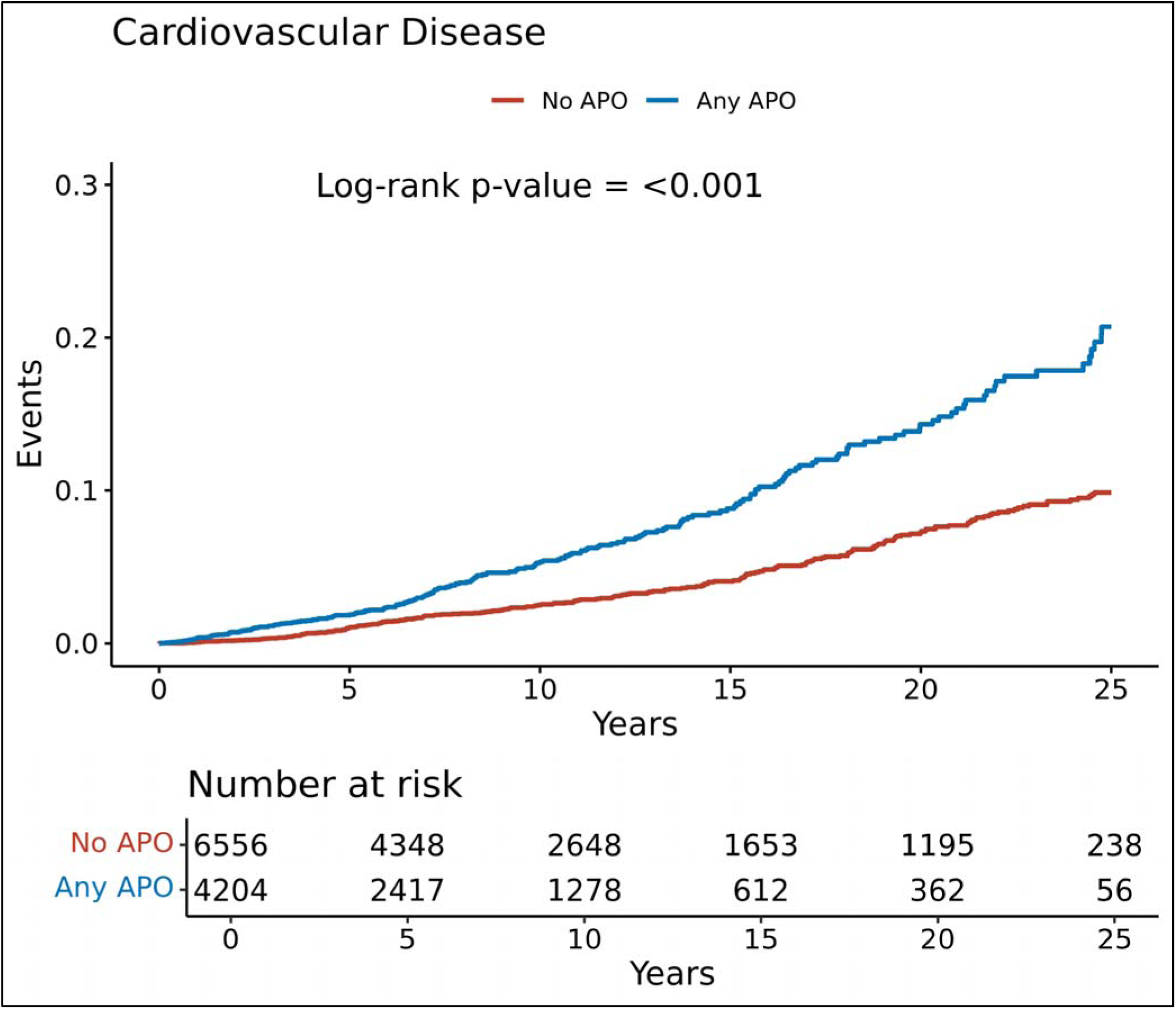
Cumulative incidence of the primary composite outcome of cardiovascular disease among women with a single recorded delivery by history of any adverse pregnancy outcome.

**Table 2.** Cox Proportional Hazard analyses results for the primary composite outcome of cardiovascular disease and the secondary outcomes of ischemic stroke, hemorrhagic stroke, myocardial infarction and heart failure.

| Outcome | Covariate Model 1<sup>a</sup> | Covariate Model 2<sup>b</sup> |
| --- | --- | --- |
|  | HR (95% CI) | HR (95% CI) |
| **Primary Outcome:** |  |  |
| Cardiovascular Disease | 1.92<br>(1.58 – 2.34) | 1.45<br>(1.17 – 1.80) |
| **Secondary Outcomes:** |  |  |
| Ischemic Stroke | 1.72<br>(1.21 – 2.45) | 1.36<br>(0.92 – 2.02) |
| Hemorrhagic Stroke | 2.32<br>(1.08 – 4.99) | 2.57<br>(1.16 – 5.69) |
| Myocardial Infarction | 3.02<br>(2.05 – 4.44) | 2.13<br>(1.39 – 3.26) |
| Heart Failure | 2.36<br>(1.82 – 3.05) | 1.64<br>(1.23 – 2.19) |

Abbreviations: HR – hazard ratio; CI – confidence interval.
<sup>a</sup>: Model 1 adjusted for age at first delivery, race, and ethnicity.
<sup>b</sup>: Model 2 adjusted for model 1 covariates + cardiovascular risk factors (hypertension, diabetes mellitus, hyperlipidemia, overweight/obese, smoking, and atrial fibrillation).

Results of interaction tests using product terms revealed that race and ethnicity significantly modified the association between APO and CVD (interaction-*p*=0.003). As shown in Figure 3, compared with White women without APO, Black women with APO had a four-fold higher risk of CVD (HR: 4.04; 95% CI: 2.81–5.79), which was also substantially greater than the risk observed in White women with APO (HR: 2.01; 95% CI: 1.43–2.84).

**Figure 3.**
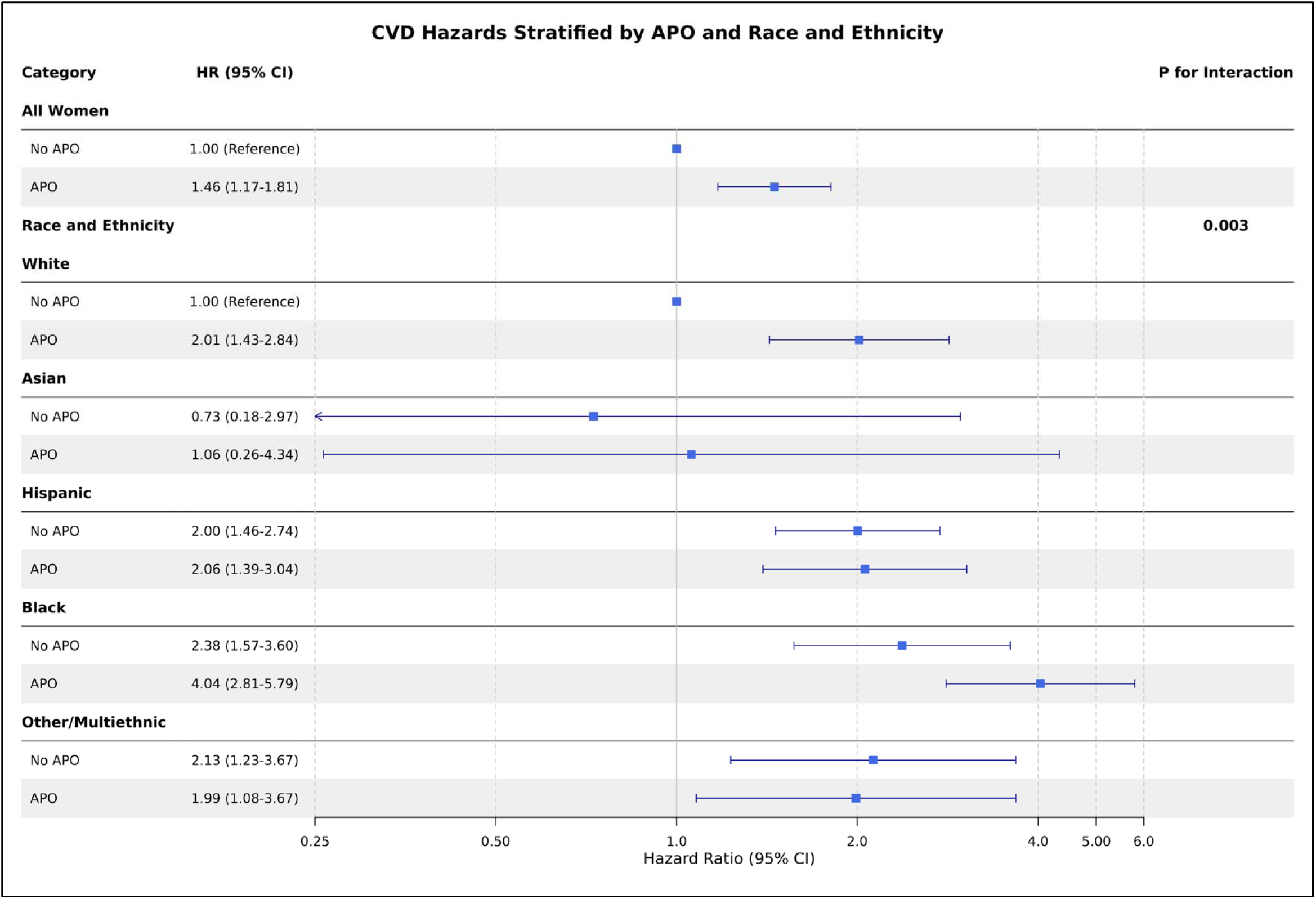
Forest plot showing the risk of cardiovascular disease by history of any APO per strata of race and ethnicity. Abbreviations: CVD – cardiovascular disease; APO – adverse pregnancy outcomes; HR – hazard ratio; CI – confidence interval.

### Secondary Analyses: Ischemic Stroke, Hemorrhagic Stroke, Myocardial Infarction and Heart Failure

Among 4,241 women with APO, 60 (1.4%) experienced an ischemic stroke, <20 a hemorrhagic stroke, 72 (1.7%) a myocardial infarction and 133 (3.2%) heart failure, while among women without APO, 73 (1.1%) experienced an ischemic stroke, <20 a hemorrhagic stroke, 44 (0.7%) a myocardial infarction and 111 (1.7%) heart failure (Figure 4, all log-rank *p*<0.01). As shown in Table 2, fully adjusted Cox proportional hazards models confirmed these results for hemorrhagic stroke, myocardial infarction and heart failure, showing that women with APO had a significantly increased hazard of each of these secondary outcomes when compared to those without APO. Ischemic stroke, however, was non-significant in the fully adjusted model (Table 2). Results were consistent across all covariate-adjusted models.

**Figure 4.**
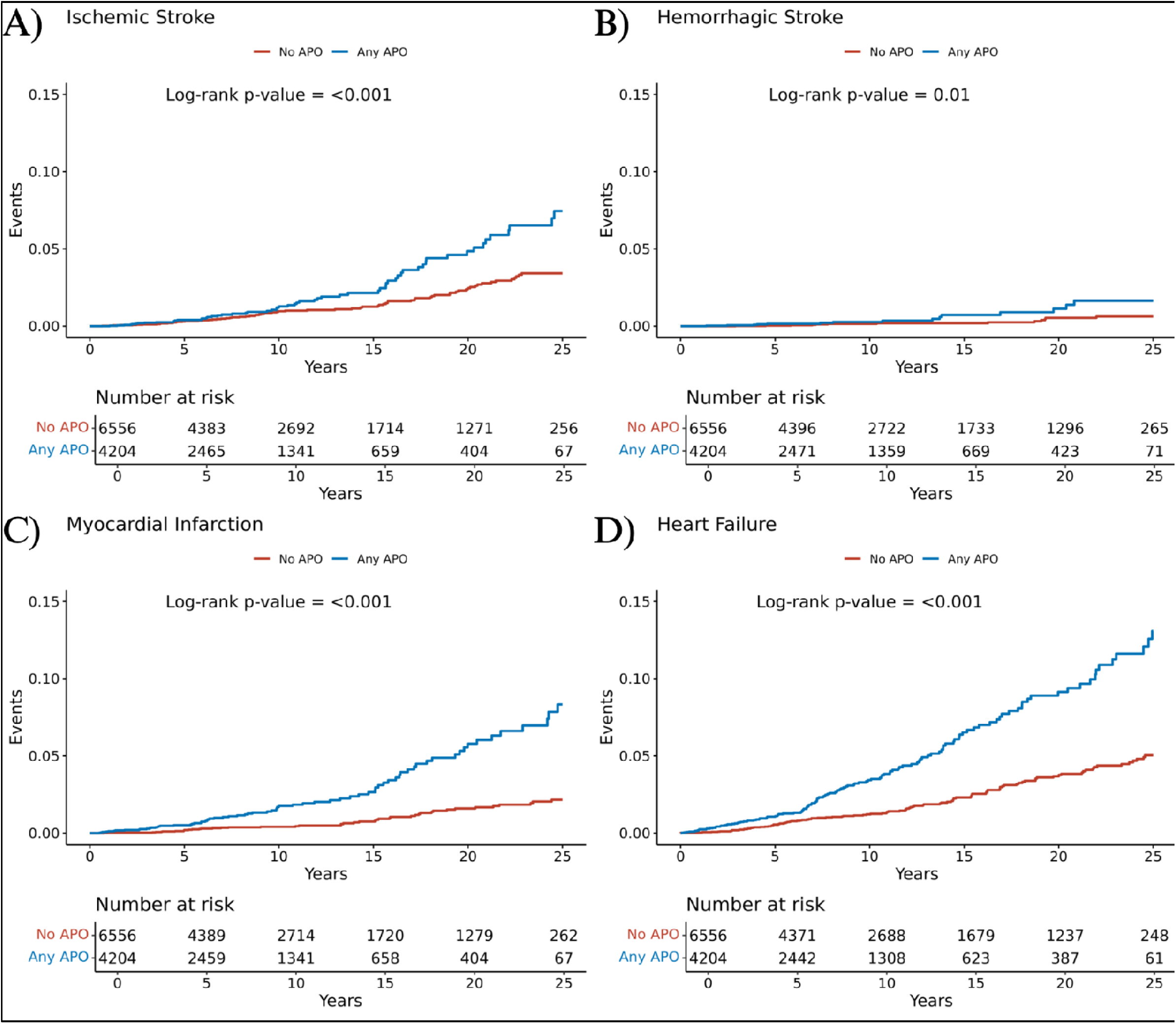
Cumulative incidences of the secondary outcomes, ischemic stroke (A), hemorrhagic stroke (B), myocardial infarction (C), and heart failure (D), among women with a single recorded delivery by history of any adverse pregnancy outcome. Created in https://BioRender.com

### Sensitivity Analyses

Findings from all three sensitivity analyses were consistent with the primary analysis. Across approaches, a history of APO remained significantly associated with higher CVD risk (effect estimates ranging from 1.45–1.52, Table 3) and the interaction between APO and race and ethnicity persisted (interaction-*p* ≤0.05, Table 3), with Black women with APO consistently showing the highest relative hazard (Table 3 and Figures S2 to S4).

**Table 3.**
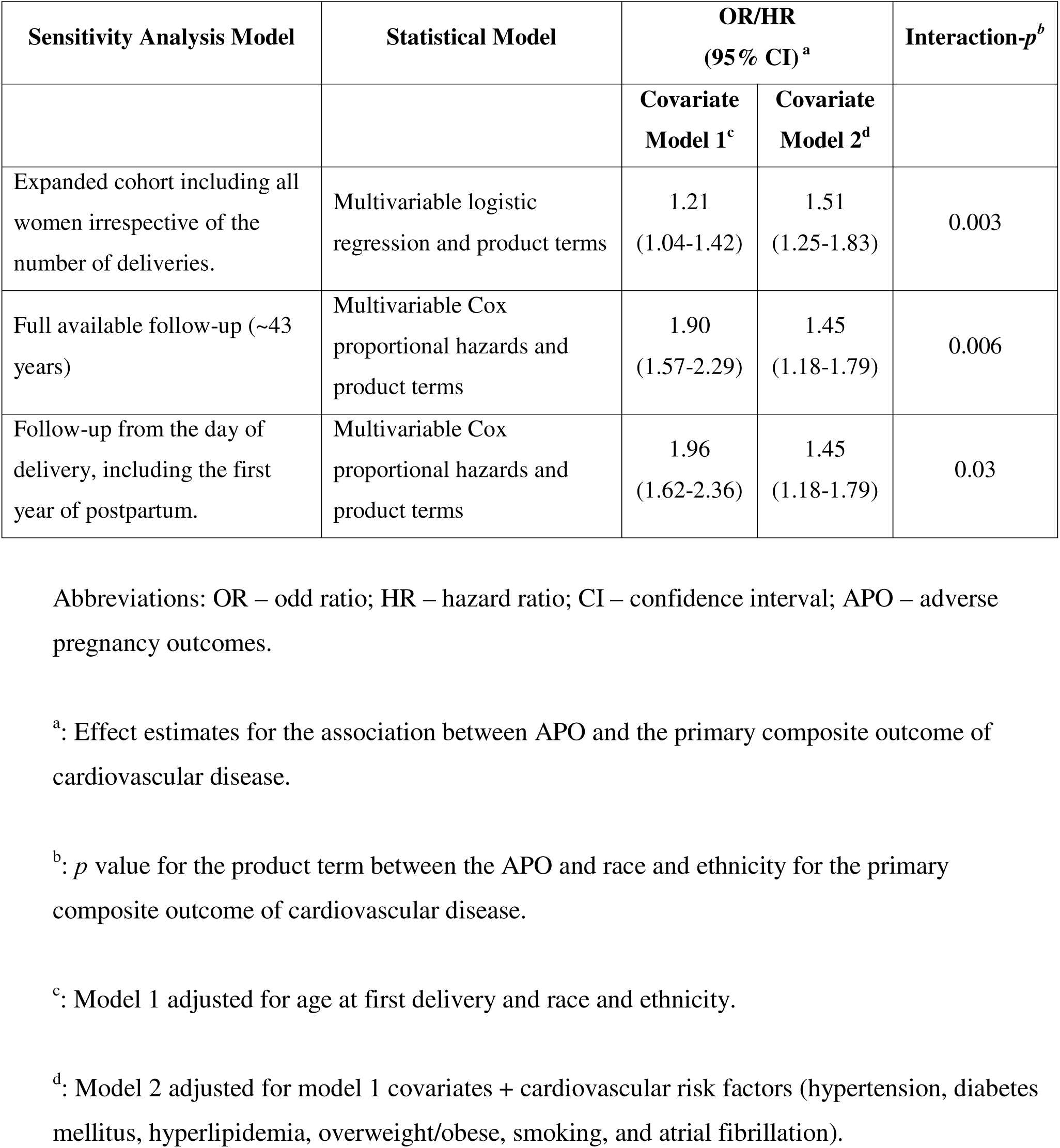
Results of sensitivity analyses.

| Sensitivity Analysis Model | Statistical Model | OR/HR<br>(95% CI)<sup>a</sup> |  | Interaction-*p*<sup>b</sup> |
| --- | --- | --- | --- | --- |
|  |  | Covariate<br>Model 1<sup>c</sup> | Covariate<br>Model 2<sup>d</sup> |  |
| Expanded cohort including all women irrespective of the number of deliveries. | Multivariable logistic regression and product terms | 1.21<br>(1.04-1.42) | 1.51<br>(1.25-1.83) | 0.003 |
| Full available follow-up (~43 years) | Multivariable Cox proportional hazards and product terms | 1.90<br>(1.57-2.29) | 1.45<br>(1.18-1.79) | 0.006 |
| Follow-up from the day of delivery, including the first year of postpartum. | Multivariable Cox proportional hazards and product terms | 1.96<br>(1.62-2.36) | 1.45<br>(1.18-1.79) | 0.03 |

Abbreviations: OR – odd ratio; HR – hazard ratio; CI – confidence interval; APO – adverse pregnancy outcomes.
<sup>a</sup>: Effect estimates for the association between APO and the primary composite outcome of cardiovascular disease.
<sup>b</sup>: *p* value for the product term between the APO and race and ethnicity for the primary composite outcome of cardiovascular disease.
<sup>c</sup>: Model 1 adjusted for age at first delivery and race and ethnicity.
<sup>d</sup>: Model 2 adjusted for model 1 covariates + cardiovascular risk factors (hypertension, diabetes mellitus, hyperlipidemia, overweight/obese, smoking, and atrial fibrillation).

### Exploratory Analyses: Age at CVD and each separate APO condition

Fully adjusted multivariable linear regressions showed that women with APO sustained CVD 2.18 years earlier than those without APO (β: −2.18; 95%CI: −3.91 to −0.45, Table 4). Additionally, multivariable Cox regressions between each separate APO condition and CVD showed that preeclampsia (HR: 1.50; 95%CI: 1.14–1.96), fetal growth restriction (HR: 1.54; 95%CI: 1.15–2.07) and gestational hypertension (HR: 1.30; 95%CI: 1–1.70) were driving the association. In contrast, preterm birth, gestational diabetes, eclampsia, and placental abruption were non-significant, although these were also the APOs with smaller numbers (Table S6).

**Table 4.** Linear regression results of APO and age at the primary composite outcome of cardiovascular disease.

| Covariate Model | Age at first cardiovascular disease event |  |
| --- | --- | --- |
|  | Beta <sup>a</sup><br>(95% CI) | *p* |
| Model 1<sup>b</sup> | -3.70<br>(-5.27 to -2.12) | <0.001 |
| Model 2<sup>c</sup> | -2.18<br>(-3.91 to -0.45) | 0.01 |

Abbreviations: CI – confidence interval.
<sup>a</sup>: Interpreted as years of earlier cardiovascular disease event.
<sup>b</sup>: Model 1 adjusted for age at first delivery, race, and ethnicity.
<sup>c</sup>: Model 2 adjusted for model 1 covariates + cardiovascular risk factors (hypertension, diabetes mellitus, hyperlipidemia, overweight/obese, smoking, and atrial fibrillation).

## DISCUSSION

In this large cohort study of diverse, parous women with one recorded delivery and without a prior history of CVD, we found that APOs were associated with a 45 percent higher risk of CVD in later life, and importantly, that this heightened risk was even greater among Black women. These results were also consistent for all parous women irrespective of their number of deliveries, using all available follow up time and including the postpartum period. In addition, we found that a history of APO was linked to a younger age at the time of CVD, and that preeclampsia, fetal growth restriction, and gestational hypertension were the APOs most strongly associated with CVD in later life.

Our findings provide important new evidence on the determinants of CVD risk in women with a prior APO. ^18,19^ Earlier studies either did not evaluate racial and ethnic differences or did not report them in their results. It is well established that non-White women face higher risks of APOs and birth complications (e.g.: low birth weight, preterm birth and infant mortality) compared to White women.^20–22^ For example, a report by Breathett et al. found that African American women had significantly higher rates of preeclampsia than White women, with prevalence odds ratios as high as 1.75 (95 % CI, 1.73 to 1.78).^20^ In a separate study of women in New York City, infants born to African American, Hispanic, and Asian mothers had a 10 to 20 percent increased risk of birth complications compared to those born to White mothers.^22^ Although prior work has shown that non-White women face higher risks of both adverse pregnancy outcomes,^20^ and CVD^23^, there is a notable lack of research examining the link between these two risks in racial and ethnic minority populations. In our study, we found that race and ethnicity significantly modified the association between APOs and MACE. Among African American women with a history of APOs, the risk of MACE was four times higher than among White women without such a history. Even when compared to White women with a history of APO, African American women had nearly twice the risk (hazard ratio, 4.04 vs 2.01, respectively).

Our study adds new insight into the relationship between APOs and CVD by showing that a history of APO is associated with an earlier onset of CVD. These findings have important implications for both primordial (i.e.: preventing the occurrence of risk factors) and primary prevention strategies, which could begin in the primary care setting or even immediately after hospital discharge following delivery.^3,11^ As our results suggest, women with a history of APO may experience CVD two to three years earlier than those without such a history. If confirmed in future studies, these findings could support the need for more proactive and timely prevention strategies in this population. For example, primary care providers may consider closer follow-up or a lower threshold for cardiovascular risk factor and disease screening in women with a history of APOs, or at least in those with APOs that were strongly associated with CVD, like preeclampsia, fetal growth restriction or gestational hypertension.

Taken together, our findings support the development, or upgrade, of cardiovascular risk assessment tools (e.g.: ACSVD or PREVENT)^24,25^ that incorporate reproductive health history, including a history of APOs, along with non-medical determinants of health such as race and ethnicity. It is essential to consider these women-specific risk factors when aiming to fully capture cardiovascular risk profiles in women.^26,27^ In 2021, the American Heart Association published a scientific statement highlighting the importance of APOs in the evaluation of CVD risk among women.^1^ That statement emphasized the lack of data on the relationship between APOs and CVD risk in diverse populations, and called for changes in health care systems to better address the unique cardiovascular needs of women.^3^ Our study contributes to this area by providing new insights from a large and diverse cohort of parous women, showing that cardiovascular risk may be especially elevated among Black women with a history of APOs. Beyond refining risk prediction, an equally critical implication of our findings is the need to ensure more intensive postpartum follow-up for women at highest risk, including earlier and more frequent screening for modifiable cardiovascular risk factors and timely initiation of preventive strategies. However, further research is needed to explore how targeted interventions, whether lifestyle-based or pharmacological, could be used to improve CVD prevention in all women, and especially in those from populations that face disproportionate risk.^28–31^

Our results also highlight important differences in CVD risk by type of APO. Consistent with previous research,^16^ preeclampsia, fetal growth restriction, and gestational hypertension were most strongly associated with CVD. In contrast, we found no significant associations for eclampsia, preterm birth, or placental abruption. However, these null findings may reflect limited statistical power from smaller sample sizes rather than the absence of a true biological relationship. For example, eclampsia, which represents the severe end of the preeclampsia spectrum, remains an important APO and cardiovascular risk factor. In our analysis, the small sample size (n=37) likely constrained power, yet the effect estimates were directionally consistent with those for preeclampsia and approached statistical significance. Regarding preterm birth and placental abruption, meta-analyses have reported associations with CVD,^16^ yet a large Norwegian cohort study of over 18,000 parous women found no link between preterm birth and cardiovascular events. Similarly, several studies have reported no association between placental abruption and later CVD.^32–34^ These discrepancies may reflect differences in population characteristics, baseline risk profiles, or follow-up duration. It is possible that some populations experience increased CVD risk from any APO, whereas in others, risk is driven primarily by specific conditions such as preeclampsia or gestational hypertension. Additionally, the timing of CVD risk may differ by APO, with some exerting greater influence in the early postpartum period and others having a more sustained impact. Future studies should examine this heterogeneity and the time-dependent nature of CVD risk by APO across diverse populations.

Our study has at least two important strengths. First, it includes a large and diverse cohort of parous women, nearly 60% of whom identified as non-White. In contrast, most prior studies in this area have examined populations that were 80–95% White.^1,18^ Second, the consistency of our findings across multiple analytical models and sensitivity analyses lends additional credibility to our results.

However, some limitations should be considered. Because delivery data were obtained through electronic health records, there is potential for misclassification, coding errors, or incomplete documentation, which could bias results toward the null. Nonetheless, the observation of strong and consistent associations despite these potential limitations increases confidence in our findings. Second, the observational nature of the study precludes causal inference, and residual confounding from unmeasured factors remains possible, even though results were robust across different covariate models and sensitivity analyses. Third, the study was underpowered to assess associations in certain racial and ethnic subgroups, such as Asian women. Finally, although we found that Black women with APOs face an especially high risk of CVD, the reasons for this disparity remain unclear. Possible contributors include differences in treatment and control of CVD risk factors, lower access to or utilization of healthcare, and broader non-medical determinants of health. Further research is needed to clarify these mechanisms and inform targeted prevention strategies.

In conclusion, our study provides novel evidence that the CVD risk associated with APOs is not uniform across populations, but that is amplified among Black women. This race and ethnicity interaction highlights the need to move beyond one-size-fits-all prevention models and toward risk assessment strategies that explicitly integrate reproductive history with the non-medical and structural determinants that shape cardiovascular health. Future research should focus on elucidating the mechanisms underlying these disparities, whether they are biological, behavioral, structural, or a combination, and on developing targeted interventions to mitigate the disproportionate burden of CVD in affected populations.

## ACKNOWLEDGEMENTS

We are grateful to the *All of Us* participants for their contributions, without whom this research would not have been possible.

