## Supplementary Figures for "Disparities in Cardiovascular Disease in Women with Adverse Pregnancy Outcomes"

*Chavers-Rivera and Clocchiatti-Tuozzo et al.*

**SUPPLEMENTARY FIGURES**

**Figure S1:** Log–log plot comparing women with and without a history of adverse pregnancy outcomes. The approximate parallelism of the curves supports the proportional hazards assumption.

**Figure S2:** Sensitivity analysis forest plot, showing hazard ratios for cardiovascular disease associated with adverse pregnancy outcomes, stratified by race and ethnicity, among all women irrespective of number of deliveries.

**Figure S3:** Sensitivity analysis forest plot, showing hazard ratios for cardiovascular disease associated with adverse pregnancy outcomes, stratified by race and ethnicity, using all available follow-up time.

**Figure S4:** Sensitivity analysis forest plot, showing hazard ratios for cardiovascular disease associated with adverse pregnancy outcomes, stratified by race and ethnicity, including events occurring during the first year postpartum.

**Figure S1.**

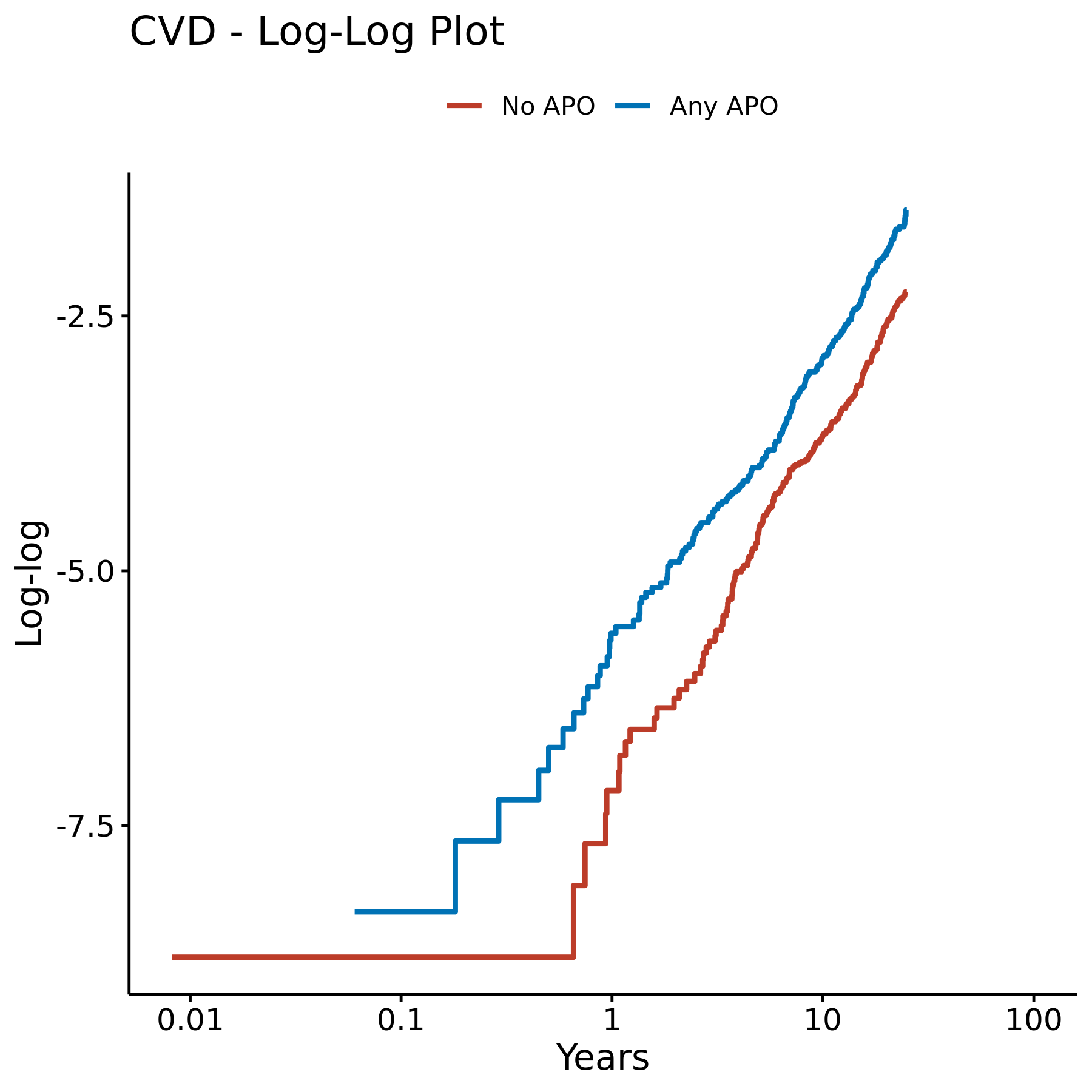

Abbreviations: CVD – cardiovascular disease.

**Figure S2.**

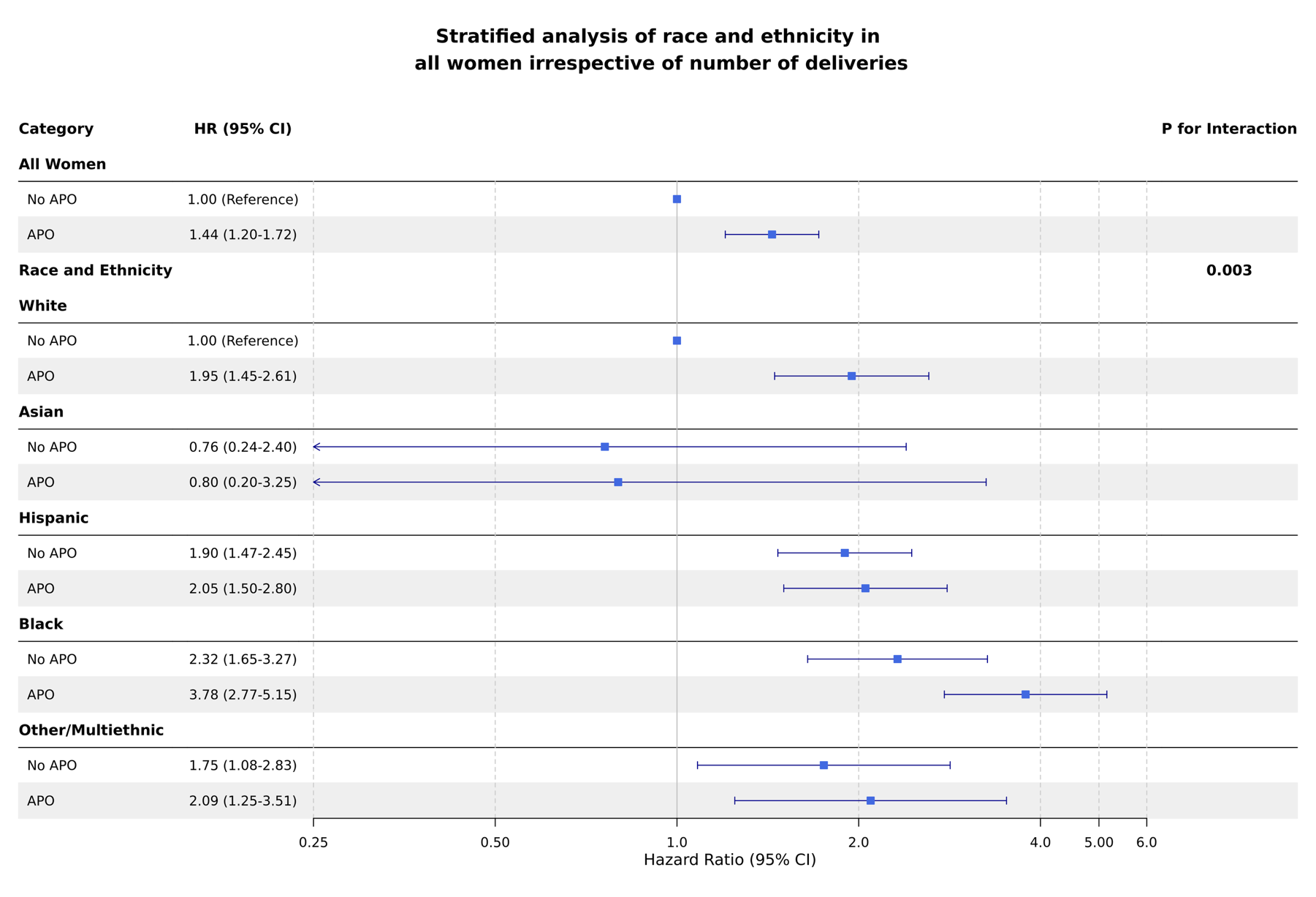

Abbreviations: HR – hazard ratio; CI – confidence interval; APO – adverse pregnancy outcome.

**Figure S3.**

Abbreviations: HR – hazard ratio; CI – confidence interval; APO – adverse pregnancy outcome.
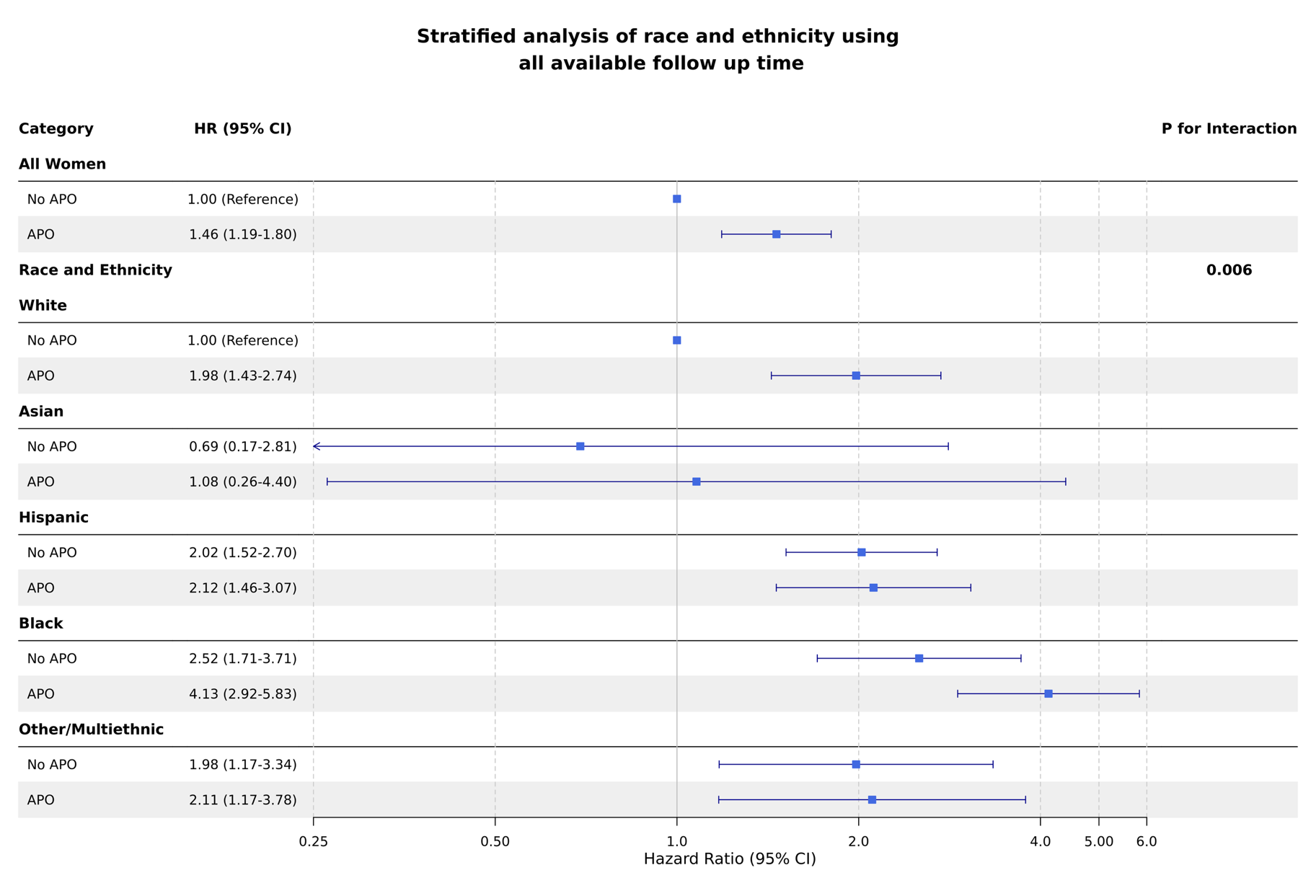

**Figure S4.**

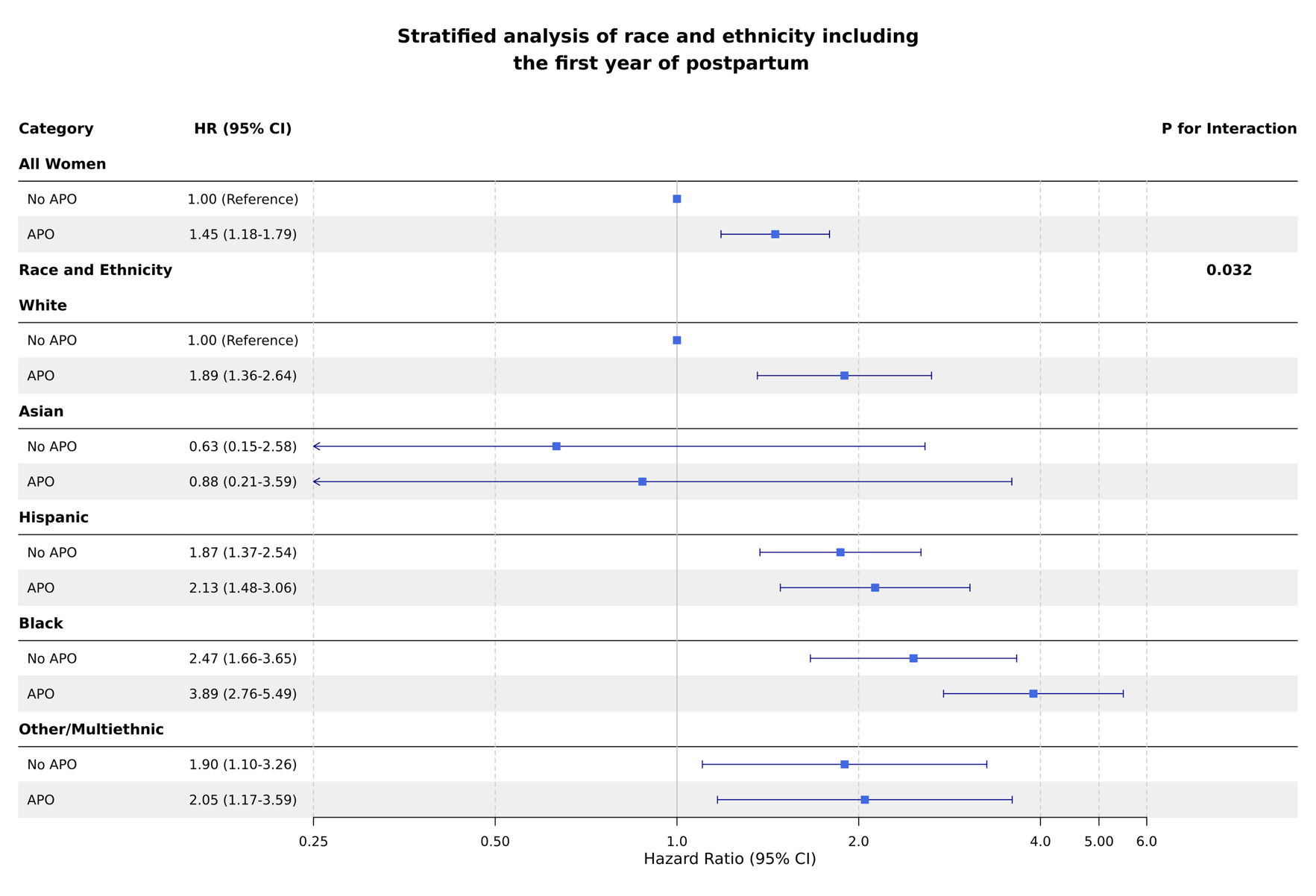

Abbreviations: HR – hazard ratio; CI – confidence interval; APO – adverse pregnancy outcome.
